# What works and for whom in treating depression in older adults in deprived communities in Brazil: Findings from a causal mediation analyses of the PROACTIVE trial that overlapped with the COVID-19 pandemic

**DOI:** 10.1101/2023.06.26.23291868

**Authors:** Nadine Seward, Carina Akemi Nakamura, Tim J Peters, Wen Wei Loh, Dean McMillan, Simon Gilbody, Marcia Scazufca, Ricardo Araya

## Abstract

**Background:** The PROACTIVE trial was a task-shared, collaborative care, psychosocial intervention that was highly effective at improving recovery from depression in older adults in Brazil that overlapped with the COVID-19 pandemic. Here we investigate mediators of the intervention’s effectiveness.

**Methods:** Causal mediation analysis using interventional indirect effects, decomposed the total effect of PROACTIVE on recovery from depression (PHQ-9<10), into multiple indirect effects including: dose of intervention (number of sessions and number of activities completed); social support measured through Luben Social Network Scale; perceived loneliness through the three-item UCLA questionnaire; conditions associated with frailty; and extra sessions offered to participants who did not respond to the intervention.

**Findings:** Of the intervention’s total effect (difference in probability of recovery from depression between the intervention and control arms 0·211 [bias-corrected 95% CI: 0·139, 0·274]): 14% was mediated through improved conditions associated with frailty 0·030 [0·003, 0·065]); 6% through reduced loneliness (0·013 [0·001, 0·028]); and 20% through attending extra sessions for participants who did not respond to the intervention (0·042 [0·007, 0·105]).

**Interpretation:** Our findings emphasise the importance of a home-based intervention to improve depression outcomes where participants are encouraged to self-select activities to mitigate against loneliness and are referred to primary care to manage health issues relating to frailty. Importantly, our findings suggest that offering extra sessions to participants who did not respond to the intervention shows promise in ensuring a sustained recovery from depression.

**Funding:** São Paulo Research Foundation and Joint Global Health Trials UK.

## Research in context

### Evidence before this study

There is a rapidly growing body of evidence demonstrating the effectiveness of behavioural activation (BA) in improving depression outcomes in older adults. However, there is little evidence on potential mechanisms through which these therapies work and for whom. Evaluating the mediators through which BA for depression operates can help to identify features of the intervention that can be optimised to improve longer-term outcomes.

Before starting these investigations, we searched PubMed for analyses evaluating potential mediators through which psychological therapies based on BA improve depression outcomes in older adults. We found only one study involving BA in older adults that took place before the pandemic. Findings of this analysis suggest that social impairment (captured using the Behavioural Activation for Depression subscale) mediated an improvement in the recovery from depression. However, this study included older patients based in Philadelphia, Pennsylvania with age-related macular degeneration that did not evaluate for the same mediators as was done in our analyses, so it is not possible to compare results.

### Added value of this study

To the best of our knowledge, we report the first robust application of the causal mediation potential outcomes-based framework of interventional indirect effects, to understand mediators through which the intervention worked (i.e., potential active ingredients). Importantly, we demonstrate that despite the fact the intervention did not specifically target issues with frailty and loneliness, both mediated an improvement in recovery from depression. Our findings also suggest benefits from attending extra sessions for participants who did not experience an early response to therapy.

### Implications of all the available evidence

Psychological therapies based on BA for older adults with depression, should consider a home-based programme delivered by community health workers that collaborates with primary care to address health issues in the older population such as those relating to frailty. Participants should also be encouraged to engage in meaningful and enjoyable activities to not only improve levels of activation, but also levels of perceived loneliness. Crucially, to improve longer-term outcomes, consideration should be given to ensuring participants attend extra sessions if they do not initially respond to therapy. To optimise interventions to support the older population, future research should focus on using robust implementation research to adapt existing progrogrammems to target the crucial ingredients identified in our analyses, whilst ensuring such solutions meets the needs and priorities of the population it serves.

## Background

Depression is the leading cause of disability globally, and a major contributor to the overall burden of disease.(1) The older population are particularly vulnerable, as depression is the most common geriatric psychiatric disorder and a major risk factor for disability and mortality.(2–4) A National Health Survey in Brazil found a higher prevalence of depression in people aged 60 or over compared with younger adults.(5) Besides the disproportional burden in the older population, lack of access to care is more common among those living in the most economically deprived areas of Brazil.(6)

The recent COVID-19 pandemic left the older population not only at greater risk of mortality and morbidity, but also at greater risk of depression through lockdowns and social distancing.(7) Even though isolation was important to controlling the pandemic and saving lives, an unintended consequence was the increased vulnerability in older people to a range of negative social, psychological, and physical health issues such as those that influence frailty. (8–11) This is especially relevant in Brazil as this was one of the worst-affected countries by the COVID-19 pandemic.(12)

The PROACTIVE study aimed to evaluate the effectiveness of a task-shared and stepped care, collaborative care psychosocial intervention delivered by community health care workers (CHWs) to improve depression recovery rates among older adults in Guarulhos, Brazil.(13) However, the COVID-19 pandemic that took hold in Brazil stopped all face-to-face intervention activities including participant recruitment and CHWs delivering sessions. Despite this, PROACTIVE was shown to be highly effective in improving recovery from depression both at the eight and at the 12-month follow-up assessments.(13)

We aim to use interventional effects to evaluate important mediators that influenced recovery from depression during a time of such adversity including: dose of the intervention (number of sessions attended and proportion of homework completed), social support, loneliness, health issues that are known to influence frailty, and the extra sessions offered to participants who did not initially respond to therapy.

## Methods

### Setting

This is a secondary data analysis using data from the PROACTIVE trial that took place in the city of Guarulhos, Brazil between May 2019 and February 2021. (14) The PROACTIVE study included 20 (Unidades Basicas de Saude (UBSs)).

### Design

PROACTIVE was a cluster randomised trial. Participants were aged 60 years or over, scored 10 or more on the Patient Health Questionnaire (PHQ-9), and were registered with one of the participating UBSs. Patients were excluded if they had: cognitive, communication, or visual problems; were unable to engage in the trial for 12 months; another person in the household included in the study; or acute suicidal risk at screening assessment. Signed informed consent was obtained before starting the screening and the baseline assessments. If the screening interview was conducted by phone, verbal consent was recorded.

### The intervention

PROACTIVE was a 17-week programme delivered by CHWs during a series of scheduled home sessions. The intervention was based on principles of psychoeducation(15) and behavioural activation.(16) Sessions were delivered by CHWs with the support of animated videos delivered via an application (app) installed on tablets.(23) Participants were educated about depressive symptoms, taught simple ways to cope with symptoms and associated problems, the importance of engaging in pleasant or meaningful activities, and relapse prevention strategies.(17–19) Some of the videos delivered as part of the sessions focused on promoting a healthy lifestyle (eating well and doing physical activities). CHWs also checked chronic conditions and helped to establish referrals to the primary care centres if necessary.

The intervention was divided into an initial phase (3 weeks) followed by a second phase (14 weeks). The first phase involved three weekly sessions focused on psychoeducation to all participants. Once the initial phase was completed, participants were assigned to either low- or high-intensity regimen focusing on behavioural activation and relapse prevention techniques. If PHQ-9 scores were assessed by the CHW as greater than or equal to 10 in session 2 and/or session 3, participants were assigned to the high-intensity regimen and received eight additional sessions. Participants assigned to the low-intensity regimen received five additional sessions.

Enhanced usual care was delivered to participants in both trial arms. UBS managers were informed about all participants included in the study. They could share information about the depressive symptomatology collected by the research assistants with members of the UBS and discuss a treatment plan as part of the enhanced usual care.

## Interruption to the intervention due to COVID-19 social distancing requirements

It is important to note that most of the sessions were delivered to participants who had been recruited before social distancing restrictions were enforced and all face-to-face activities ended.(13) The main limitation with regards to this mediation analyses is that the majority of follow-up visits were conducted over the phone and not face-to-face as originally intended. As a consequence, we were not able to collect information for the Behavioural Activation for Depression Scale - Short Form that required visual aids.

### Measures

#### Exposure

Participants in the intervention arm were offered PROACTIVE intervention (exposed) whereas participants in the control arm were offered enhanced usual care (unexposed).

#### Outcome

We used the secondary outcome of recovery from depression measured at 12 months and defined as PHQ-9 scores less than 10.

#### Mediators

A mediators is defined as a variable that is on the causal pathway between the initial exposure to either intervention or control, and the outcome (recovery from depression measured at 12 months). It is therefore a variable that is causally influenced by the intervention and in turn causally influences the outcome.(20) A full description of the mediations and selection criterion can be found in the supplementary material. The following mediators selected for these analyses include:

### Measures to capture the dose of the intervention (M1)

We anticipated that the ‘dose’ of the intervention would not only influence whether participants recovered from depression, but also other mediators on the pathway to recovery from depression. To capture this effect, we created a mediator to reflect the number of sessions (M1a: 0-8 sessions) attended by participants that was independent of the extra sessions attended by participants who did not respond to the intervention. We also created another mediator to reflect the number of activities a participant completed (M1b). We categorised the proportion of assigned activities completed as follows: no assigned activities; 1-5 assigned activities; and 6-13 assigned activities.

### Social support (M2)

We theorised that the intervention that was based on behavioural activation would have encouraged participants to engage with, and seek support from, family and friends and hence reduce social isolation. To capture this effect, we used the six-item Lubben Social Network Scale (LSNS) at eight months. Higher LSNS scores indicate greater levels of perceived social support.

### Loneliness (M3)

We theorised that during the COVID-19 pandemic, the PROACTIVE intervention that encouraged engagement in meaningful or enjoyable activities, could improve recovery from depression through a reduction in loneliness. The UCLA three-item loneliness score (M3a) was used to capture levels of perceived loneliness.(21) Scores ranged from 3 to 9, where higher scores indicated increasing levels of loneliness. Follow-up assessments that took place after social distancing requirements took hold due to the COVID-19 pandemic, also collected data on the impact of the pandemic on whether participants felt more alone as a result of the pandemic (M3b; 0=no; 1=yes).

### Frailty (M4)

Frailty is has been defined as a complex geriatric syndrome which could be influenced by a range of symptoms including pain, mobility and balance problems, and weakness; all of which can lead to depression.(22) During lockdown, it was reported that the older population were particularly vulnerable to increased levels of frailty, that was strongly associated with depression.(23) We theorised that PROACTIVE could have improved symptoms that influenced frailty that in turn improved depression outcomes. There were two features of the PROACTIVE intervention that theoretically could have improved levels of frailty. Firstly, given CHWs helped participants access primary care for any health issues they were experiencing (collaborative care component of the intervention), it is possible that conditions associated with frailty improved, which facilitated the recovery from depression. It is also possible that the sessions that encouraged participants to engage in a healthy lifestyle by encouraging them to be more active, could have improved levels of frailty. We captured this potential mediator using various conditions that PROACTIVE could theoretically improve including postural imbalance (M4a) and use of a crutch (M4b). It is worth noting that these were the only two conditions associated with frailty that we were able to capture. Due to the face to face sessions being cancelled, all measures of chronic health conditions that could influence frailty were not possible.

### Extra sessions received in instances of non-response to the intervention (M5)

Adding extra sessions for participants who do not respond to the intervention by sessions two or three, may help to improve symptoms of depression. Therefore, if a participant did not respond to the intervention by session two or three (PHQ-9≥10), they were offered the high-intensity regimen with three additional sessions. Estimating this indirect effect via the additional sessions involves two variables including non-response to treatment (M5a: 0=non-response; 1=responded to treatment) and the number of extra sessions received (M5b: 0=no extra sessions; one to three extra sessions).

### Statistical methods

#### General

To better understand the relationship between different mediators and recovery from depression, we compared different mediators and confounders with the outcome recovery from depression at 12 months (determined by a PHQ-9 score less than 10) for participants in the intervention arm only. Differences in baseline characteristics between treatment arms can be found in previous publications.(17)

#### Mediation analysis

We aimed to investigate the extent to which recovery from depression measured at 12 months using the PHQ-9 questionnaire (PHQ-9 scores less than 10) were explained by the direct and indirect effects of the intervention (Figure 1). To achieve these objectives, we used the interventional (in)direct effects approach to mediation analysis to understand population-level effects relevant to this analysis.(24) Interventional effects are the latest in a series of developments that use advanced causal inference frameworks that overcome limitations of other approaches to mediation.(25, 26) Findings for this analyses are reported according to guidelines for reporting mediation analyses (AGReMA statement).(20) Further details on the mediation analysis relating to the interventional effects can be found in the supplementary material.

**Figure 1:**
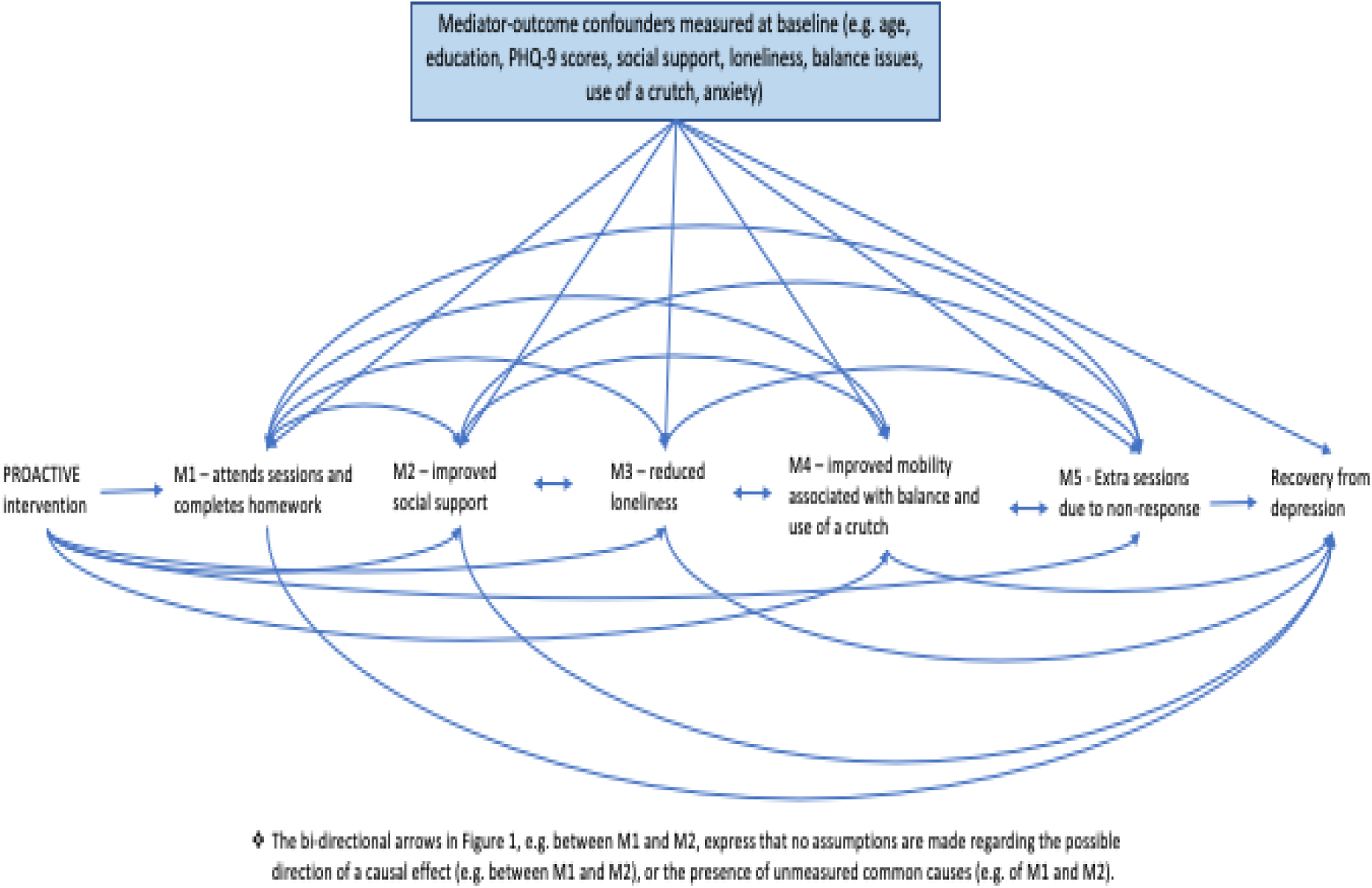
Causal model demonstrating the proposed pathways through which the PROACTIVE intervention may improve remission from depression

### Estimation and model fit

Estimation for the interventional indirect effects was based on Monte Carlo integration using a 1,000-fold expanded dataset.(24) The expanded dataset was created in four steps. Details of each step, and details of the models including interactions and selected confounders can be found in supplementary material. Bias-corrected confidence intervals were based on nonparametric bootstrap with 1,000 resamples.(24) The bootstrap also accounted for clustering at the primary care centre. Stata code using a similar approach can be found elsewhere.(26)

### Assumptions

The interventional effects have important underlying assumptions that will influence the validity of our findings if violated.(24) Due to the randomised nature of PROACTIVE, the assumptions about no unmeasured confounders between the treatment arm and each mediator, and between the treatment arm and the outcome, are fulfilled.(24) The main assumption relevant to our study is that there are no unmeasured mediator-outcome confounders.

### Sensitivity analysis

Primary outcomes for depression are mainly reported as either recovery from depression (PHQ-9 <10) or an improvement of at least 50% in PHQ-9 scores between baseline and follow-up. We selected our outcome measure of recovery from depression (PHQ-9 scores <10) to ensure consistency with the outcomes reported in the main trial paper. To facilitate comparability with other studies, we also conducted the same analyses using the outcome of improved symptoms of depression (50% reduction in PHQ-9 scores between baseline and the 12-month follow-up).

### Missing data

To account for missing data, we implemented single stochastic imputation using chained equations with 10 burn-in iterations, under the assumption that data were ‘missing at random’ (MAR). In each of the 1000 bootstrap samples, the imputation is done once and separately in the control and intervention arms. All imputation models included the mediators and confounders used in the models to assess interventional effects.

### Ethical approval and consent

This study was approved by the Ethics Committee of Universidade de São Paulo Medical School (CEP FMUSP number 2.836.569) and authorised by the Guarulhos Health Secretary. Written or, when applicable, oral informed consent to participate was obtained from all individuals before screening and baseline assessments.(17, 18)

### Role of the funding source

The MRC had no involvement in the study design, collection, analysis and interpretation of the data, in the writing of the paper, and in the decision to submit the paper for publication.

## Results

Table 1 compares mediators, in the intervention arm only, between participants who recovered from depression (PHQ-9 scores below 10), and those with scores at or above this point using complete data only. Without adjustment for mediator-outcome confounders, these descriptive statistics indicate that participants who received a higher dose of the intervention (M1a, M1b) had a small increased probability at recovering from depression at 12 months. There was little evidence to support the association between LSNS scores captured at eight months (M2) and recovery from depression at 12 months. Participants who recovered from depression at 12 months had lower levels of perceived loneliness captured using the UCLA three-item questionnaire (M3a) at eight months than participants who did not recover. Participants who did not have any issues that with postural imbalance at eight months were more likely to recover from depression at 12 months (M4a). There was not much difference in crutch use between participants who recovered from depression at 12 months and those participants who remained depressed (particularly after adjusting for crutch use at baseline). Lastly, participants who did not experience an early response to therapy (M5a) and attended extra sessions in instances of non-response (M5b), were more likely to recover from depression at 12 months.

**Table 1:**
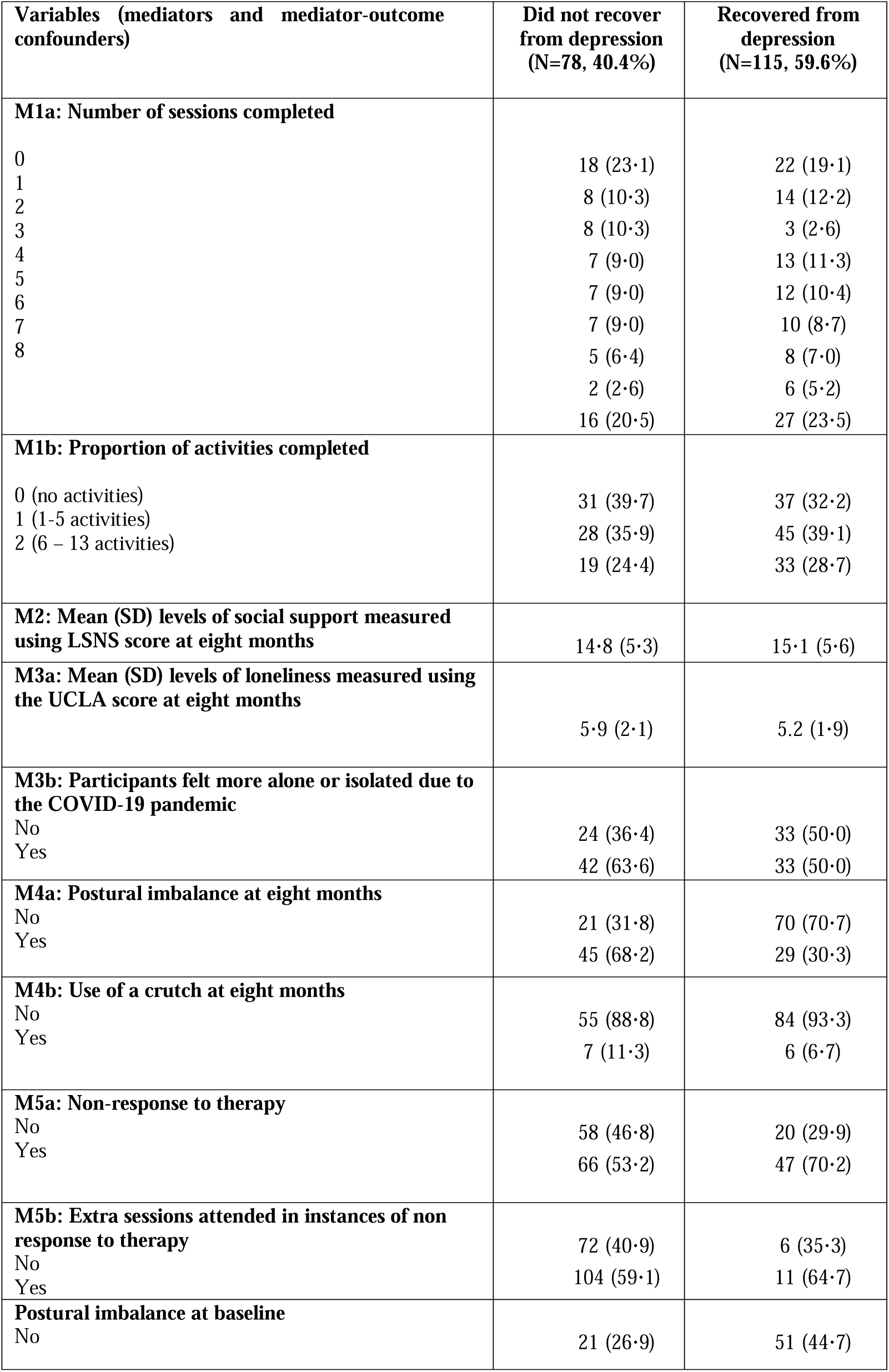
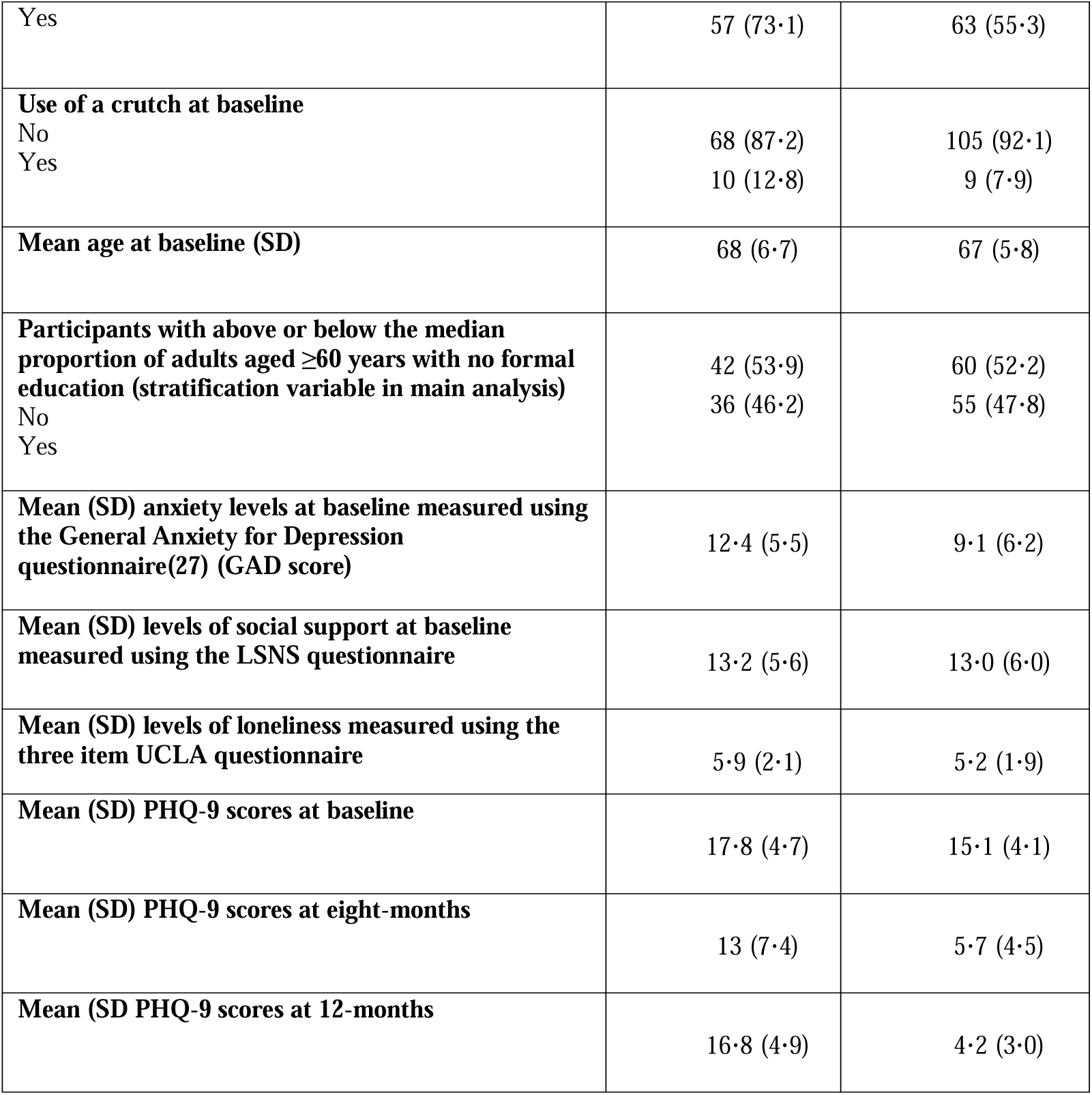
Comparison of mediators and mediator-confounders between participants with PHQ-9 scores below 10 at 12 months and those with PHQ-9 scores 10 and above (complete cases only)

### Mediation analyses

Table 2 demonstrates that at 12 months the difference in probability of recovery from depression between the intervention and control arms (adjusted difference in probability of recovery from depression: 0·211, bias-corrected 95% CI: 0·139, 0·274) approximately 40% was mediated through our selected mediators. Specifically, 14% of the total effect was mediated through improved health issues relating to frailty (M4a, postural imbalance and M4b, use of a crutch) at eight months (adjusted difference in probability of recovery from depression via improved physical health: 0·030, bias-corrected 95% CI: 0·003, 0·065). Furthermore, 6% was mediated through a reduction in loneliness (adjusted difference in probability of recovery from depression via UCLA scores: 0·013, bias-corrected 95% CI: 0·001, 0·028). The additional sessions (M5a) offered to non-responders (M5b) mediated 20% of the total effect in recovery from depression (adjusted difference in probability of recovery from depression via extra sessions for non-responder: 0·042, bias-corrected 95% CI: 0·007, 0·105)

**Table 2.** Total effect and interventional (in) direct effects of the PROACTIVE intervention on recovery from depression (PHQ-9<=10) at 12 months follow-up

| Effects | Estimates<br>(bias-corrected 95% CI)<sup>a,b,c</sup> |
| --- | --- |
| Total effect of PROACTIVE on improved symptoms of depression | 0.211 (0.140, 0.274) |
| Direct effect | 0.090 (0.047, 0.198) |
| Indirect effect of attending sessions and completing assigned activities (M1) | 0.035 (-0.034, 0.125) |
| Indirect effect of social support (M2) | -0.011 (-0.027, 0.001) |
| Indirect through loneliness (M3) | 0.013 (0.001, 0.028) |
| Indirect effect through improved health issues influencing frailty (M4) | 0.030 (0.003, 0.065) |
| Indirect effect through additional sessions for non-responders (M5) | 0.042 (0.006, 0.105) |

<sup>a</sup> Estimates have been adjusted for mediator-outcome confounders of age, education, baseline PHQ-9 scores, baseline UCLA scores, baseline LSNS scores, baseline GAD scores, baseline issues with postural imbalance, baseline use of a crutch.
<sup>b</sup> Estimation for the different effects was based on Monte Carlo integration using 1,000-fold expanded dataset
<sup>c</sup> Bias-corrected confidence intervals were based on nonparametric bootstrap with 1000 resamples adjusting for clustering

### Sensitivity analyses

Estimates from our analyses using as outcome the outcome measure of a reduction in PHQ-9 scores of at least 50% between baseline and the sixth month follow-up, were largely consistent with our findings that used the outcome of recovery from depression (PHQ-9 >=10. Findings are reported in the supplementary information.

## Discussion

Using interventional indirect effects, the causal mediation analyses provide insights into how the PROACTIVE intervention achieved an effect in the recovery from depression at 12 months. Whilst simultaneously accounting for all mediators, we have been able to demonstrate the importance of targeting health issues relating to frailty in the older population in the recovery from depression. Improved levels of loneliness at eight months, also facilitated recovery from depression at 12 months. Lastly, attendance for extra sessions by initial non-responders mediated the largest improvement in recovery of depression.

The older population are particularly vulnerable to increasing levels of frailty leaving them susceptible to a range physical and mental health conditions including depression; all of which were exacerbated during the COVID-19 pandemic.(23, 28) The PROACTIVE intervention had many features that theoretically could have influenced factors associated with frailty. Firstly, the sessions delivered by the digitally supported app, encouraged participants to maintain a healthy lifestyle (being physically active, having a healthier diet, and avoiding accidents in the home), attend scheduled health appointments, and adhere to prescribed medications. Another feature of PROACTIVE was the referral of participants to primary care for their physical health needs. Although we are not able to disentangle which aspect of the PROACTIVE intervention influenced postural imbalance or use of a crutch, a likely explanation is that it was a combination of many features of the intervention. Future trials should give consideration to ensuring an older participants’ physical health, as well as frailty, is considered alongside any mental health issues.

There is growing global health concern regarding the role of loneliness in a myriad of both physical and mental health conditions.(29) There is also a small but rapidly expanding body of evidence about the role of behavioural activation in mitigating against loneliness in socially isolated older adults.(30, 31) Our results add to this evidence-base by demonstrating that an intervention based on behavioural activation led to reduced levels of loneliness, in turn facilitating recovery from depression at 12 months. Although the PROACTIVE intervention did not specifically target participants to select activities to improve levels of loneliness, it is has been suggested that interventions that target maladaptive social cognition, such as cognitive behavioural therapy, are effective at improving levels of loneliness.(32) It therefore seems reasonable to assume that behavioural activation that encourages participants to engage in enjoyable activities, may in turn facilitate improvements in loneliness. More research is needed to explore how interventions can target loneliness to improve outcomes of depression and physical health outcomes.(32)

A feature of the PROACTIVE intervention is that participants who did not respond to the therapy by session two or three were offered three additional sessions. The additional sessions offered to the participants reinforced previous session content. To account for this, we created an additional mediator to understand if the stepped care inherent in the offer of these additional sessions improved recovery from depression for participants who did not respond to therapy. Estimates suggest that the indirect effect mediated through the stepped care component of PROACTIVE, was responsible for a substantial increase in the recovery from depression. This suggests that stepped-care is an important feature of programmes to treat depression in older adults, in similar contexts.

It is difficult to ascertain exactly how the pandemic influenced findings from the PROACTIVE trial. The majority of the follow-up assessments were conducted over the phone as social distancing requirements had been enforced and all face-to-face activities stopped. Therefore, mediators captured at the eight-month follow-up assessment may have been influenced by the pandemic. It is possible that the results from our analyses that are taken from a trial that took place at times of extreme adversity, providing useful insight into important public health concerns in older adults that otherwise would not have been possible.

### Comparison with previous studies

There was one only one analyses that evaluated for potential mediators for an intervention based on behavioural activation in older adults. (33) However, this analysis evaluated for different mediators and therefore is not comparable. It is anticipated that, as findings from research begin to emerge from this rapidly expanding discipline, we will have greater insight into different mediators that help to improve recovery from depression in the older population.

Although there have been no studies on the dose of the intervention (number of sessions completed or activities completed) mediating the recovery from depression in older adults, findings from studies with different populations were similar to our findings. (34, 35) However, contrary to another analyses looking at the mediating role of extra sessions offered to adults with severe depression that did not experience an early response to the sessions, we did find evidence to support the benefit of the ‘stepped-care’ feature, in the recovery from depression.(26) More research is needed to better understand for whom and in what contexts stepped-care interventions improve symptoms of depression.

### Strengths and limitations

Our approach to mediation has several strengths. Importantly, our mediation analyses allowed us to simultaneously investigate multiple mediators, their interactions and non-linearities. Failing to account for the relationships between the different mediators could over- or underestimate the indirect and direct effects.

We have reported limitations of applying interventional effects to psychological therapies elsewhere.(26) In particular, for mediators that are measured in the intervention arm only (e.g.,number of sessions and homework completed) there is a possibility of non-compliance bias. There is a chance that we were not able to adjust for this with the available confounders (uncontrolled confounding). There is also a limitation whereby if we were not able to capture a mediator that influences the mediators included in our model, then our estimates could be biased. As an example, we only able capture two health issues relating to frailty that were also associated with loneliness and depression. It is possible that there are other health issues that we did not have data on, that influenced not only frailty but also the other mediators.

## Conclusions

Our robust mediation analysis has helped to better understand how a collaborative care, psychosocial intervention was able to have such a pronounced impact on the recovery from depression. Our results suggest that the PROACTIVE intervention shows promise in improving not only recovery from depression but also a myriad of other closely connected health conditions in the older population. Since loneliness is a known risk factor for both mental and physical conditions, future research should consider incorporating this as an explicit feature in psychological therapies.

## Supporting information

Supplementary materia

## Data Availability

Data available upon request

## Contributors

N.S., R.A., and M.S conceptualised the idea for the study. N.S. designed and conducted mediation analyses and wrote the initial and subsequent draft of the paper. C.N., assisted with the analyses. W.L. and T.P provided statistical guidance. N.S., R.A., M.S., C.N., W.W.L, T.P., D.M., S.G., read and edited all drafts of the paper. N.S. and C.N. has directly accessed and verified the underlying data.

## Data sharing

Upon request and subject to review, we will provide the data that support the findings of this study.

## Declarations of interest

None exist

## Acknowledgments

The original PROACTIVE study was funded by the Medical Research Council. We are also grateful for the participants who agreed to be involved with this study.

## Notes

### Competing Interest Statement

The authors have declared no competing interest.

### Clinical Trial

ISRCTN57805470

### Funding Statement

Sao Paulo Research Foundation and Joint Global Health Trials UK

### Author Declarations

This study was approved by the Ethics Committee of Universidade de Sao Paulo Medical School (CEP FMUSP number 2.836.569) and authorised by the Guarulhos Health Secretary

