## Supplementary materia for "What works and for whom in treating depression in older adults in deprived communities in Brazil: Findings from a causal mediation analyses of the PROACTIVE trial that overlapped with the COVID-19 pandemic"

**(160+403+18)**

**Supplementary material**

1. **Selection of potential mediators**

We included two types of mediators: variables that were measured in the intervention arm only; and variables that were measured in both the intervention and control arms. Mediators measured only in the intervention arm are variables that are a direct consequence of receiving the PROACTIVE intervention and therefore not measured for participants in the control arm. Mediators measured in the intervention arm only were included in the final model if they were associated with another mediator, or the outcome (*p*<0.10). Potential mediators measured in both arms of the trial were only included in the final model if they were associated with the exposure (*p*<0·10) or the outcome (*p*<0·10). Using the above criteria for our two types of mediators, we can capture any potential mediator that is statistically influenced by the intervention, and influences either another mediator, or the improvement in depressive symptoms. Failing to include putative mediators that are concurrently confounders of mediator-outcome relations for other mediators can cause severe bias. For this reason, we elected to err on the side of caution and include mediators empirically associated with the outcome (or another mediator) to reduce the chances of confounding bias; hence the relatively large p-value threshold.

A causal mediation analysis compares mediators in the exposed population, to the unexposed population (i.e., the counterfactual), while fixing the values of the remaining mediators under a specific exposure level.(20) For example, the direct effect of the intervention compares the potential improvement in depression symptoms between the intervention and control, while holding loneliness fixed at values found in the control arm (in other words, their counterfactual values). The fundamental problem of causal inference rules out observing counterfactual mediator values under both intervention and control for each individual. Therefore, these must be imputed using the observed data. The counterfactual values of the unexposed levels of the mediators measured in both arms of the trial are estimated using observed values from participants in the control arm. Conversely, exposed levels of the mediators are estimated using observed values from participants in the intervention arm. The values of mediators measured in the intervention arm only were categorised so that an ‘unexposed’ value of zero was defined for those who were assigned to the intervention arm but were not exposed to the mediator of interest (such as those who did not attend any sessions). In other words, we assumed that after adjusting for relevant confounders, participants in the control arm who could not have accessed the PROACTIVE intervention, had the same unexposed value of zero as participants in the intervention arm who could have been but in practice were not exposed to the mediator of interest. Stata code available in a previous publication helps to conceptualise how this was achieved in practice.

2. **Selection of mediator-outcome confounders**

Due to the randomised nature of participants being allocated to the intervention or control arm, the exposure, it was not necessary to consider confounding for the association between treatment allocation and the outcome, or between the treatment allocation and the mediators. However, it was necessary to account for confounders potentially distorting the association between the mediator and the outcome (mediator-outcome confounders). We considered all demographic characteristics as potential mediator-outcome confounders.

Mediator-outcome confounders are variables that influence the outcome or any one of the mediators (p<0.10) and included the following variables captured at baseline: age (continuous), PHQ-9 scores, participants with above or below the median proportion of adults aged ≥60 years with no formal education (yes/no), anxiety (GAD-7 assessment scores(30), social networking (LSNS scores), perceived loneliness (UCLA scores), postural imbalance, and use of a crutch. There were interactions between the following mediator-outcome confounders: GAD scores and LSNS scores, education and age, and education and UCLA scores.

3. **Mediation analyses**

*Decomposition of total effect of the PROACTIVE intervention into direct and indirect effects*

We decomposed the total effect of the PROACTIVE intervention into the interventional direct effect and the indirect effects via each of the five mediators. For the decomposition to be valid, the sum of the direct and indirect effects must be the same as the total effect of the PROACTIVE intervention.(23)

*Estimation and model fit*

Estimation for the interventional indirect effects was based on Monte Carlo integration using a 1,000-fold expanded dataset. (15) The expanded dataset was created in four steps. In the first step we fitted a model for each mediator given exposure and other predictors. Specifically, we fitted linear (M1a, M2, M3a, M3b), ordinal (M1b), and logistic (M4a, M4b, M5a, M5b) regression models to the observed data. Each model included a combination of predictors that were shown to be associated with the mediator of interest (*p*<0.10) including: age, education, baseline PHQ-9 scores, baseline LSNS scores, baseline UCLA scores, use of a crutch or postural imbalance issues at baseline, and gender. Interactions and non-linearities were explored and included if they had *p*<0.10 using Stata’s post-estimation command, *testparm*.

In the second step, the fitted mediator models were used to generate random, subject-specific Monte Carlo draws of each mediator for both the exposed and unexposed condition (the latter being the counterfactual), given their observed covariate values. For mediators measured in both arms of the trial, exposed status refers to allocation to the intervention arm, and unexposed status refers to allocation to the control arm. For mediators measured in the intervention arm, exposed refers to being exposed to mediator of interest (attended sessions vs did not attend sessions; completing homework vs not completing homework; response to treatment vs did not respond; attended extra sessions vs did not attend any extra sessions).

In the third step, we fitted a model for the outcome using a logistic regression model for recovery from depression, separately in the exposed and unexposed groups, given the mediators and mediator-outcome confounders. Any potential mediator-outcome confounder was included if it was associated with the mediator or the outcome (*p*<0·10). We used a model selection criterion similar to that of the mediator models – that is, any relevant non-linearities and interactions with *p*<0·10 were included in the outcome model, using the post-estimation *testparm* command in Stata. All non-linearities and interactions can be found in Supplement 1.

There were important mediator-mediator interactions between UCLA scores (M3) and participants who attended extra sessions (M5). Specifically, participants with high levels of loneliness at eight months (M3), were more likely to recover from depression if they attended extra sessions (M5b), compared with participants with high levels of loneliness who did not respond to the sessions and who did not attend extra sessions.

In the fourth step, we used the fitted outcome model to predict the potential outcomes in the expanded dataset given the random, subject-specific draws of the mediator counterfactuals from the second step. The interventional indirect effects were then calculated as the mean differences between potential outcomes under different hypothetical exposure levels.

**4. Sensitivity analyses**

Primary outcomes for depression are mainly reported as either recovery from depression (PHQ-9 <10) or an improvement of at least 50% in PHQ-9 scores between baseline and follow-up. We selected our outcome measure of recovery from depression (PHQ-9 scores <10) to ensure consistency with the outcomes reported in the main trial paper. To facilitate comparability with other studies, we also conducted the same analyses using the outcome of improved symptoms of depression (50% reduction in PHQ-9 scores between baseline and the 12-month follow-up) to ensure consistency with the outcomes reported in the main trial.

**Table 1: Sensitivity analyses comparing estimates from interventional indirect effects between the outcome of recovery from depression at six-months and reduction in PHQ-9 scores by 50% between baseline and six-months in the pooled analyses, the Lima trial, and the trial in São Paulo**

**Table 1: Pooled analysis (n=1045)**

| **Effect** | **Reduction in PHQ-9 scores by 50% between baseline and sixth months** | **Recovery from depression at six months (PHQ-9 scores less than 10)** |
| --- | --- | --- |
|  | **Estimates (bias-corrected 95% CI)^a,b,c^** | **Estimates (bias-corrected 95% CI)^a,b,c^** |
| Total effect of PROACTIVE on improved symptoms of depression | 0.232 (0·157, 0·300) | 0·211 (0·140, 0·274) |
| Direct effect | 0·131 (0·082, 0·254) | 0·090 (0·047, 0·198) |
| Indirect effect of attending sessions and completing assigned activities (M1) | 0·007 (-0·074, 0·122) | 0·035 (-0·034, 0·125) |
| Indirect effect of social support (M2) | 0·005 (-0·009, 0·027) | -0·011 (-0·027, 0·001) |
| Indirect through loneliness (M3) | 0·027 (0·012, 0·052) | 0·013 (0·001, 0·028) |
| Indirect effect through improved health issues associated with frailty (M4) | 0·022 (0·005, 0·045) | 0·030 (0·003 0·065) |
| Indirect effect through additional sessions for non-responders (M5) | 0·042 (-0·004, 0·105) | 0·042 (0·006, 0·105) |

^a^ Estimates have been adjusted for mediator-outcome confounders of age, education, baseline PHQ-9 scores, baseline UCLA scores, baseline LSNS scores, baseline GAD scores, baseline issues with balance, baseline use of a crutch.

^b^ Estimation for the different effects was based on Monte Carlo integration using 1,000-fold expanded dataset

^c^ Bias-corrected confidence intervals were based on nonparametric bootstrap with 1000 resamples adjusting for clustering

**_**
